# Characterization of *de novo* structural variants in trios with orofacial clefts

**DOI:** 10.64898/2026.10.16.26365772

**Authors:** Kelsey R. Robinson, Sarah W. Curtis, Nehir E. Kurtas, Alba Sanchis-Juan, Alexandra Manchel, Wasiu Lanre Adeyemo, Terri H. Beaty, Azeez Butali, Carmen J. Buxó, David J. Cutler, Michael P. Epstein, Eleanor Feingold, Lord JJ Gowans, Jacqueline T. Hecht, Lina Moreno Uribe, Carmencita D. Padilla, Gary M. Shaw, Seth M. Weinberg, Jeffrey C. Murray, Justin Cotney, Michael Talkowski, Mary L. Marazita, Harrison Brand, Elizabeth J. Leslie-Clarkson

## Abstract

Structural variants (SVs) are larger genetic rearrangements such as deletions, duplications, and insertions and other forms of structural alterations. Given their large size, they can have an outsized effect on phenotype, however, the contribution of SVs to the genetic architecture of orofacial clefts (OFCs) is not well understood. In this study, we sought to better understand the contribution SVs to OFC risk by characterizing *de novo* SVs (dnSVs) in 1,549 probands with an OFC. Overall, we called 375 dnSVs in 307 probands (0.242 dnSVs per genome). The dnSV rate per genome was highest among individuals with cleft lip (CL, 0.29), followed by cleft lip and palate (CLP, 0.25) and cleft palate only (CP, 0.20), though differences across subtypes were not statistically significant. There were also no differences between sex or syndromic status, but we did identify a significant difference in the rate of duplications across OFC subtypes (p=0.005). Compared to CP (0.028), duplication rates were significantly higher for both CL (0.121; p = 0.003) and CLP (0.096; p = 0.002), suggesting a potential etiologic mechanism for CL/P. We systematically curated each dnSV to classify its potential for influencing OFC risk by examining the genes and regulatory elements disrupted by the SV and evidence for their role in craniofacial development or OFCs. We identified 22 dnSVs in regions associated with recurrent genomic disorders, including seven deletions and one duplication of the well-established OFC risk locus 22q11.2. Additionally, we identified 12 large (>1Mb) SVs, with eight meeting criteria for a likely pathogenic classification based on total gene involvement, and seven dnSVs disrupting well-known genes associated with autosomal dominant forms of OFC. The most notable was *SATB2,* for which we found four dnSVs, all in individuals with CP. Next, by combining evidence from recurrent dnSVs and *de novo* single nucleotide variants with Hi-C contact maps and predicted chromatin states from developing craniofacial tissues, we found new candidate genes for OFC risk including *FHAD1*, *NBEA*, *NR3C2*, and *SPHKAP*. Overall, dnSVs classified as likely pathogenic accounted for putative molecular diagnoses in approximately 1.67% of the cohort, with an additional 0.96% having variants of unknown significance with plausible risk for OFCs. Our results support dnSVs as an important component of OFC etiology and suggest their inclusion in analyses may improve diagnostic yield for affected individuals.

## Introduction

Structural variants (SVs) are DNA rearrangements including copy number variants such as deletions and duplications, as well as inversions, translocations, and complex chromosomal rearrangements^1^. They are by definition at least 50bp in size and can range up to many megabases of DNA. These rearrangements can influence gene dosage through gain or loss of DNA or regulatory elements, or result in position effects in which the rearrangement impacts linear or three-dimensional chromatin structure without changes to dosage^2, 3^. Copy number changes have long been implicated in human disease since 1965 when the 4p-(Wolf-Hirschhorn Syndrome) was described^4, 5^, as these types of rearrangements are detectable by a variety of technologies^6^. As such, there are currently 66 established copy number-associated syndromes in DECIPHER^7^ (accessed 05/05/26). Still, with the advent of short- and long-read sequencing technologies, larger and more detailed catalogues of human structural variation are now possible. As sequencing has been adopted for discovery and clinical use, the contribution of SVs to all types to human disease has been refined. Although it was known that large rearrangements detected by karyotype or microarray explain ∼35% of congenital anomalies, smaller events detected by sequencing are estimated to account for 2-8% of congenital anomalies in a variety of cohorts^8–12^.

Among congenital anomalies, orofacial clefts (OFCs) occur at a rate of 1 in 1000 live births, making them the most common craniofacial birth defect. There are three main subtypes of OFCs: cleft lip (CL), cleft lip and palate (CLP), and cleft palate (CP), though OFCs involving the upper lip are often combined into a single group of CL with or without CP (CL/P). Approximately 30% of CL/P and 50% of CP occurrences are accompanied by other structural or developmental anomalies and are considered syndromic. Many syndromic forms of OFCs are thought to have a monogenic etiology caused by large chromosomal rearrangements or single nucleotide variants in the coding sequence. Historically, isolated or non-syndromic OFCs were thought to have a complex etiology reflecting a multifactorial load of genetic and environmental risk factors. However, due to variable expressivity, incomplete phenotyping, and the increased use of sequencing in genetic studies, a role for rare pathogenic variants—often in genes associated with syndromes—has become clear, blurring etiologic distinction between syndromic and non-syndromic OFCs ^13–18^.

SVs are known to play a role in both syndromic and non-syndromic OFCs. For example, SVs associated with syndromic OFCs include 22q11.2 deletions (DiGeorge syndrome, MIM: 188400)^19^, 1p36 deletions (MIM: 607872)^20^, 4p deletions (Wolf-Hirschhorn syndrome, MIM: 194190)^21, 22^, and 17p11.2 deletion (Smith-Magenis syndrome, MIM: 182290)^23^. Smaller microdeletions have also been reported for *IRF6*^24^, *TFAP2A*^25^, *TP63*^26^, and other genes in which exonic single-nucleotide variants cause syndromic OFCs. Similarly, evidence of a role for SVs in non-syndromic OFCs comes from studies of copy number variants^27–30^, including deletion of *TP63* with variable expressivity within the same family. Some early studies used array-CGH and deviations from Mendelian inheritances on microarrays to identify deletions and duplications in individuals with OFCs^31, 32^. Larger studies using SNP array data from genome-wide association studies have found evidence for association of *de novo* deletions on 7p14.1 with OFCs^27, 33^ and inherited deletions in *MGAM*, *ADAM3A*, and *ADAM5*^34^. A larger array-CGH study of 1,102 individuals by Lansdon *et al*^30^ included both syndromic and non-syndromic probands with CL/P and CP. They found no difference in SV burden between any group comparison but did describe three new OFC candidate genes—*COBLL1*, *RIC1*, and *ARHGEF38*.

Because the higher resolution of whole-genome sequencing (WGS) data allows for detection of a range of SVs in terms of size and SV type, we aimed to expand the understanding of SVs in OFC risk by investigating *de novo* structural variants (dnSVs) in a WGS dataset of 1,549 trios with OFCs and mixed syndromic status. Smaller *de novo* single nucleotide variants (dnSNVs) have been found to be enriched in cohorts of both CL/P^14, 35^ and CP^15^, providing insight into candidate genes for OFCs; therefore, we hypothesized dnSVs found in probands with OFCs could also lead to gene discovery.

## Methods

### Cohort Description

This study included 1,549 case-parent trios with CL (n=215), CP (n=450), or CLP (n=884) who were sequenced as part of either the Gabriella Miller Kids First Pediatric Research Program (GMKF) (n=1093 trios) or separate sequencing projects focusing on CP (n=456 trios) (Supplemental Table 1). Participants were recruited globally, reflecting that OFCs affect people of many different ancestries. This dataset included participants with European ancestry (recruited from Spain, Turkey, Hungary, United States), Latin America (Puerto Rico, Argentina), Asia (China, Singapore, Taiwan, the Philippines), and Africa (Nigeria, Ghana). Participant recruitment was done at regional treatment or research centers after review and approval by each site’s institutional review board (IRB) and the IRB of the affiliated US institutions (*e.g.,* the University of Iowa, the University of Pittsburgh, and Johns Hopkins University). Due to the varied protocols and ages of participant recruitment, it is not possible to definitively classify probands as having isolated or non-isolated OFCs. Based on the information available at the time of recruitment, most participants were classified as having an isolated cleft. However, due to the uncertainties and practical challenges of comprehensively phenotyping study participants in a cross-sectional study design, those participants with additional clinical features consistent with a syndromic diagnosis were not excluded from this analysis and comprise 3.48% (n=54) of the cohort.

### SV Calling and *De Novo* Filtering

SV detection was performed using GATK-SV, a method designed to identify, genotype and resolve all classes of SVs detectable in whole-genome sequencing (WGS)^36^. This includes both balanced and unbalanced copy number variants (CNVs), inversions, insertions, translocations, and 11 types of complex SVs. Briefly, GATK-SV maximizes sensitivity by integrating five distinct algorithms and subsequently adjudicates and re-genotypes SVs based on raw read evidence^37^. GATK-SV integrates all SV signature evidence available from WGS, including discordant paired-end (PE), split reads (SR), normalized read-depth (RD) and B-allele frequencies (BAF). Each CRAM file undergoes processing with five algorithms: two PE/SR-based methods (Manta, Wham)^38, 39^, two RD-based methods (cnMOPS, GATK-gCNV)^40, 41^, and a mobile element detection algorithm, MELT^42^. GATK-SV is openly accessible on GitHub (https://github.com/broadinstitute/gatk-sv) and can be deployed on the Google Cloud Platform using the Cromwell execution engine, enabling parallel scaling. Once variant discovery is complete, the resulting VCF file is initially annotated with the GATK SVAnnotate^7^ tool to assess the functional impact of SVs and compare allele frequencies against population reference databases such as gnomAD SV^37^.

*De novo* SV filtering was performed as previously described in Belyeu *et al*^43^. To start, multi-allelic CNVs and variants with unresolved breakpoints were excluded. Only variants present in the proband and absent in both parents were considered *de novo*. Additional filtering was applied as follows: 1) low-quality variants (genotype quality = 1 and called exclusively by the Wham algorithm) were removed, 2) variants overlapping regions with known somatic mutations and poor coverage (*e.g.*, immunoglobulin heavy-chain locus and Histocompatibility Complex locus) were excluded, 3) small deletions and duplications (<1000 bp) supported only by SR evidence were removed unless they had ‘both-sides support’, 4) depth-only calls smaller than 10Kbp were filtered out, and 5) deletions supported solely by PE evidence with a read depth copy number of 2 were excluded. Additionally, *de novo* highly complex SVs, mosaic variants and reciprocal translocations were manually reviewed to confirm accuracy as in Werling *et al*^44^. All SVs were visually confirmed using the IGV and Flipbook^45^ tools.

### Annotation

Genomic disorder (GD) regions, defined as disease-associated recurrent CNVs caused by non-allelic homologous recombination (NAHR), were inspected for 89 loci, 30 of which were evaluated for both deletions and duplications^46^. Both raw depth-based calls and VCF calls were assessed for overlap with these regions using bedtools intersect, requiring a reciprocal overlap of at least 30% between each call and the GD regions. For raw depth-based calls, we additionally collected scattered CNVs that covered at least 30% of the corresponding GD region using bedtools coverage. We excluded variants that showed ≥30% coverage of a GD region but failed to meet a reciprocal overlap threshold of ≥30% in bedtools intersect, thereby removing asymmetric overlaps likely driven by large CNVs spanning GD regions without a desired reciprocal support. Samples with an excessive number of raw-depth and VCF variants were removed, identified using the IQR outlier method and exceeding the upper threshold (75th percentile + 1.5 × IQR). Normalized depth plots for each GD region were manually inspected and low-quality loci were removed. Trio-based depth patterns were used to infer mode of inheritance.

For the remaining dnSVs, we used the Variant Effect Predictor (VEP, v115)^47^ to annotate overlaps with common SVs in gnomAD (v4.1) for deletions, duplications, and insertions in our dataset. We filtered gnomAD SVs to only include variants with a “PASS” filter flag, matched by variant type, and required at least a 50% intersection to consider a variant overlapping. We then used a minor allele frequency (MAF) filter of 0.1% to filter variants from our OFC dataset (excluding known genomic disordered regions), only retaining variants without common variant overlaps.

We classified variants based on a variety of factors (Supplemental Figure 1) including size, complexity, location in established genomic disorder regions, and whether they did or did not overlap a coding region. In addition, we investigated SVs that overlapped any of the 418 genes on a curated OFC gene panel^48^, or genes with *de novo* single nucleotide variation in OFC trios^15, 35^ (Supplemental Table 2). For non-coding variants, we categorized SVs that overlapped regions with potential function in the developing face to identify novel candidate genes for OFCs. For this, we evaluated RNA-seq data from human embryonic craniofacial tissue at Carnegie Stage (CS) 13-22^49^ and looked at dnSVs that overlap genes in the top 80^th^ percentile of expression or overlap putative human craniofacial enhancers^50^ from epigenomic data. For SVs that overlapped craniofacial enhancers, we also assessed whether the enhancer was located within the same topologically associated domain as nearby genes in both cranial neural crest cells (CNCC)^50^ or embryonic stem cells (ESCs)^51^. We identified recurrent dnSVs by grouping those that overlapped within 500kb upstream or downstream of each other (maximum of 1Mb apart).

### Rare inherited variants

For structural variants overlapping genes on the OFC gene panel associated with autosomal recessive (AR) modes of inheritance^13^, we searched for a second, potentially pathogenic variant in the sequencing data. The sequencing and quality control of this data has been extensively described previously^14, 52, 53^. Briefly, variants were called and aligned to the GRCh38/hg38 reference genome using either the GATK pipeline^54–56^ or the DRAGEN Germline version 3.7.5 pipeline on the Illumina BaseSpace Sequence Hub platform. Annotation of these variants was done using the Variant Effect Predictor (VEP v115)^47^.

### Statistical analyses

We compared the rate of dnSVs across several categories. For comparisons involving three groups (*e.g.,* OFC subtypes), we used the non-parametric Kruskal-Wallis test to compare the median values for number of SVs and SV length. For follow up pairwise comparison or comparison of two groups, we used a Mann-Whitney U test.

Module scores were generated from human snRNA-seq expression data^57^ using the AddModuleScore function in Seurat (v. 5.2.1) for the 16 candidate genes with dnSVs and SVs and 2 established OFC genes (*SATB2* and *IRF6*) and visualized in the form of a violin plot to identify cell subtype specific patterns. A clustered dot plot was also generated using these 18 genes to visualize the underlying gene expression patterns across the human cell subtypes. Cell subtypes were annotated based on DenovolyzeR^2^ (v. 0.2.0) enrichment (Benjamini-Hochberg corrected p-value < 0.1) of protein damaging de novo variants (DNs) in orofacial cleft trios from the Gabriella Miller Kids First program^3,4^ as previously computed in Khouri-Farah et al. 2026^1^. We converted the Seurat snRNA-seq expression data from Khouri-Farah *et al.* 2026^1^ to a CellTypeData set with the Expression Weighted Celltype Enrichment (EWCE) R package^5^ (v. 1.12.0) and then assessed linear positive correlation of cell type gene expression specificity to all 1391 genes reported to have a protein-altering dnSNVs from Supplemental Table 2. Standard deviations from the mean of bootstrapping tests performed by EWCE were used for plotting and significant enrichments were computed as Benjamini-Hochberg corrected p-value < 0.05.

## Results

### Summary of dnSVs

We evaluated a total of 1,549 trios with OFCs for dnSVs. We identified 375 dnSVs in 307 probands (Supplemental Table 3), with an average rate of 0.242 dnSVs per genome (Figure 1A), which is similar to a projected and empirical mutation rate of ∼0.29 dnSVs per generation^36, 44, 58^. Stratified by SV type, there were 158 deletions (ranging in size from 51bp to 14.7Mb) and 124 duplications (57bp to 5.6Mb). We called 64 insertions, of which 50 were transposable elements, as well as 4 balanced translocations, 1 unbalanced translocation, 2 inversions (103bp and 3.0Mb), and 22 complex rearrangements (Figure 1B).

**Figure 1:**
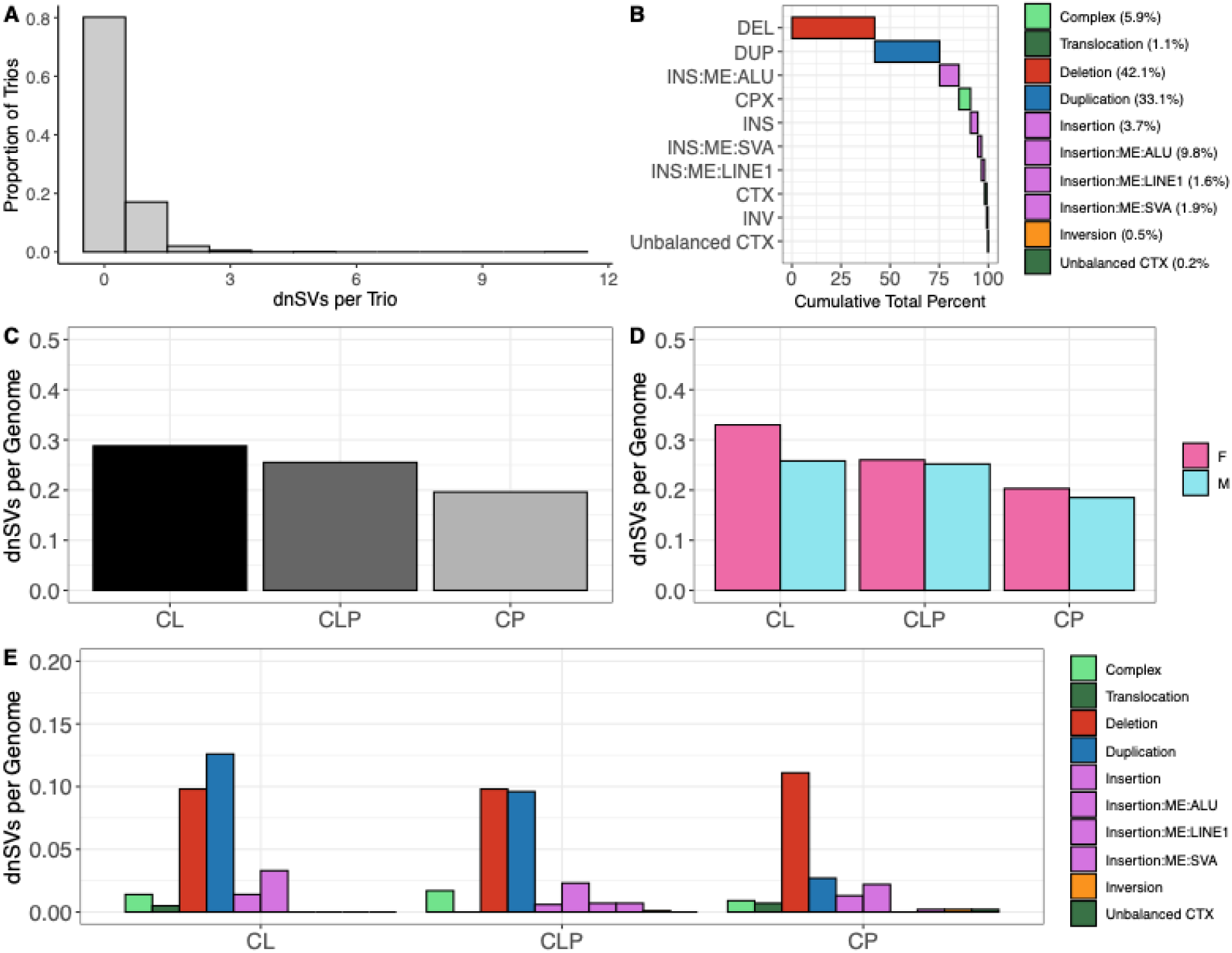
Distribution of 375 dnSVs in 1549 trios with OFCs. A) variants per proband B) classification of SVs by type C) rate of dnSVs per genome by proband OFC subtype, D) rate of dnSVs per genome by proband OFC subtype and proband sex E) rate of dnSVs per genome by OFC subtype and dnSV type

We found no difference in the dnSV rate (*i.e*., dnSVs per genome) when split by sex (male=0.238, female=0.246; p=0.63) or syndromic status (syndromic=0.247, non-syndromic=0.203; p=0.74). We note that out of 1,033 individuals with reported syndromic status, only 54 were classified as syndromic, limiting our ability to identify statistical differences by status. When split by OFC subtype, we found the rate of dnSVs was higher in CL (0.29), followed by CLP (0.25), then CP (0.20) (Figure 1C), but this difference across groups was not statistically significant (p=0.24). Because there are sex biases in the prevalence of different OFC subtypes, we repeated these comparisons stratified by OFC subtype and proband sex. The rates between male and female were not significantly different for any category: CL (male=0.27, female=0.34; p=0.94), CLP (male=0.25, female=0.216 p=0.48), and CP (male=0.19, female=0.20; p=0.79) (Figure 1D).

We next compared the type of dnSVs among OFC subtypes to determine if a specific variant type was more likely to be present in one over another. Due to sample sizes, statistical comparisons were restricted to deletions and duplications (Figure 1E). We first evaluated deletions, which occurred at similar rates across all OFC subtypes (CL=0.098, CLP=0.098, CP=0.111; p=0.74). In contrast, when we compared duplications, we found the rate was significantly different between subtypes (CL=0.121, CLP=0.096, CP=0.028; p=0.005). Further, although deletion rates were similar across all subtypes and consistent with rates in the literature (0.146 deletions per genome), the duplication rate in CL/P was notably higher than the reported rate of 0.040 per genome^36^. To further characterize these differences, we identified the highest pTriplo score^46^ in genes affected by duplications or LOEUF scores^59^ for genes affected by deletions for SVs in non-genomic disordered regions to see if the predicted impacts were more or less severe by phenotype. Although genes within CL duplications tended to have higher pTriplo scores on average (0.51), this difference was not significantly different (p=0.72) from CLP (0.40) or CP (0.41), and we observed no differences in average LOEUF scores for deletions (p>0.24; Supplemental Figure 2). Together, this data suggests *de novo* duplications may contribute to CL/P risk, though the mechanism is unclear.

Reasoning that larger SVs might be associated with more severe phenotypes, we asked whether the lengths of deletions and duplications differed by OFC subtype, sex, or syndromic status. There were no significant differences between the dnSV deletion length and OFC type (Figure 2A). However, when stratified by sex, we found females had significantly larger deletions than males (female median=6kb, male median=2.2kb; p=0.01) (Figure 2B). We observed a similar trend within each OFC subtype, but the differences in deletion length between females and males were not statistically significant (Figure 2B). As females are less likely to have CLP than males, finding that females had larger deletions could be consistent with the notion that these SVs have stronger effects, similar to the “female protective effect” suggested in autism spectrum disorder^60^.

**Figure 2:**
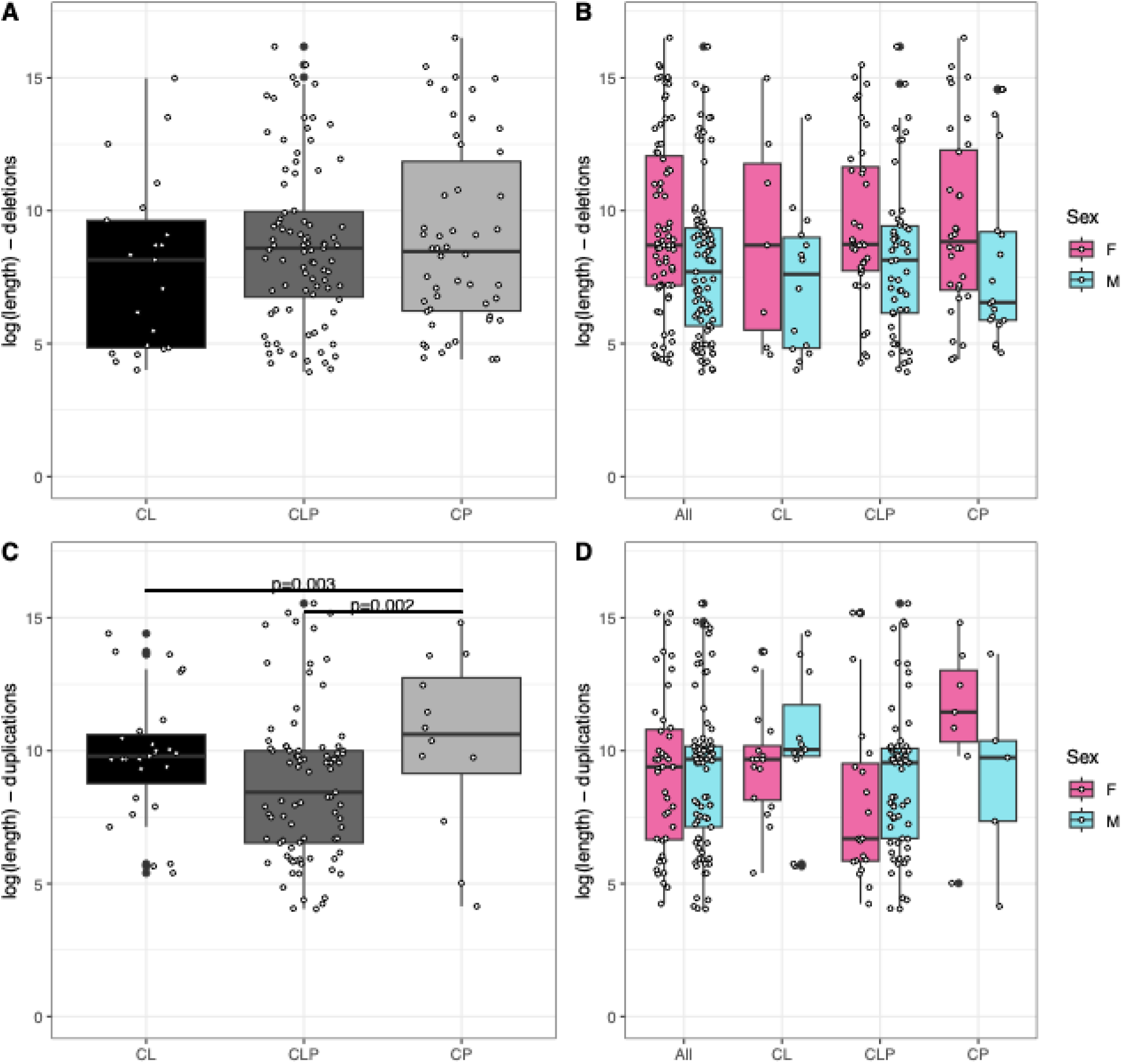
Comparison of dnSV length by SV type, OFC subtype, and proband sex. Boxplots for A) deletion length by proband OFC subtype B) deletion length by OFC subtype and proband sex C) duplication length by proband OFC subtype, D) duplication length by OFC subtype and proband sex

For duplications, we found that individuals with CLP had shorter duplications (median=4.6kb) than individuals with CL (median=18kb) or CP (median=42kb) (Figure 2C; p = 0.04). We observed no difference in duplication length males versus females (Figure 2D). In contrast to what we observed for deletions (where females consistently have higher median SV length) the median duplication length for CL and CLP was higher in males compared to females, but higher in CP females compared to CP males (Figure 2D). None of these comparisons were statistically significant, potentially due to lower power with multiple stratifications of the dataset. We did not perform statistical analysis for syndromic versus non-syndromic individuals, as there were only 4 deletions (size range 276bp to 10.5Mb) and 4 duplications (size range 63bp to 790kb).

To determine how the dnSVs might contribute to OFC risk, we systematically categorized dnSVs based on SV type, length, and the genes or regulatory elements impacted using a sequential categorization approach. Each category and subcategory is labeled in Supplemental Figure 1, which will be used throughout our results (*e.g.,* flowchart (FC) 1A for variants >1Mb in size affecting OFC-associated genes). In the following sections, we describe functional consequences of dnSVs and highlight the major findings from this exercise.

### Pathogenic dnSVs account for 1.7% of OFCs

We first asked what fraction of OFC trios could be attributed to a dnSV. For this calculation, we only considered dnSVs that are clearly interpretable according to ACMG standards^61^ and included recurrent genomic disorder regions, large SVs, and variants clearly affecting established OFC genes (Table 1). We identified 22 structural variants in regions consistent with recurrent genomic disorders (FC GDR), all of which have been associated with OFCs and/or craniofacial dysmorphisms as part of their phenotypic spectra. The most frequently identified regions were 22q11.2^62^ (7 deletions, 1 duplication) and 15q11.2^63, 64^ (3 deletions, 2 duplications). In total, if we assume all 22 variants identified in these genomic disordered regions are pathogenic and causal for OFCs, these variants account for a molecular diagnosis in 1.4% of sequenced probands. However, reduced penetrance has been noted for several of these regions, so a more conservative estimate only including highly penetrant or OFC specific variants such as 22q11.2^65^ (both deletions and the duplication), 7q11.23^66^, and 3q29^67^ is closer to 0.7% (n=11).

**Table 1:**
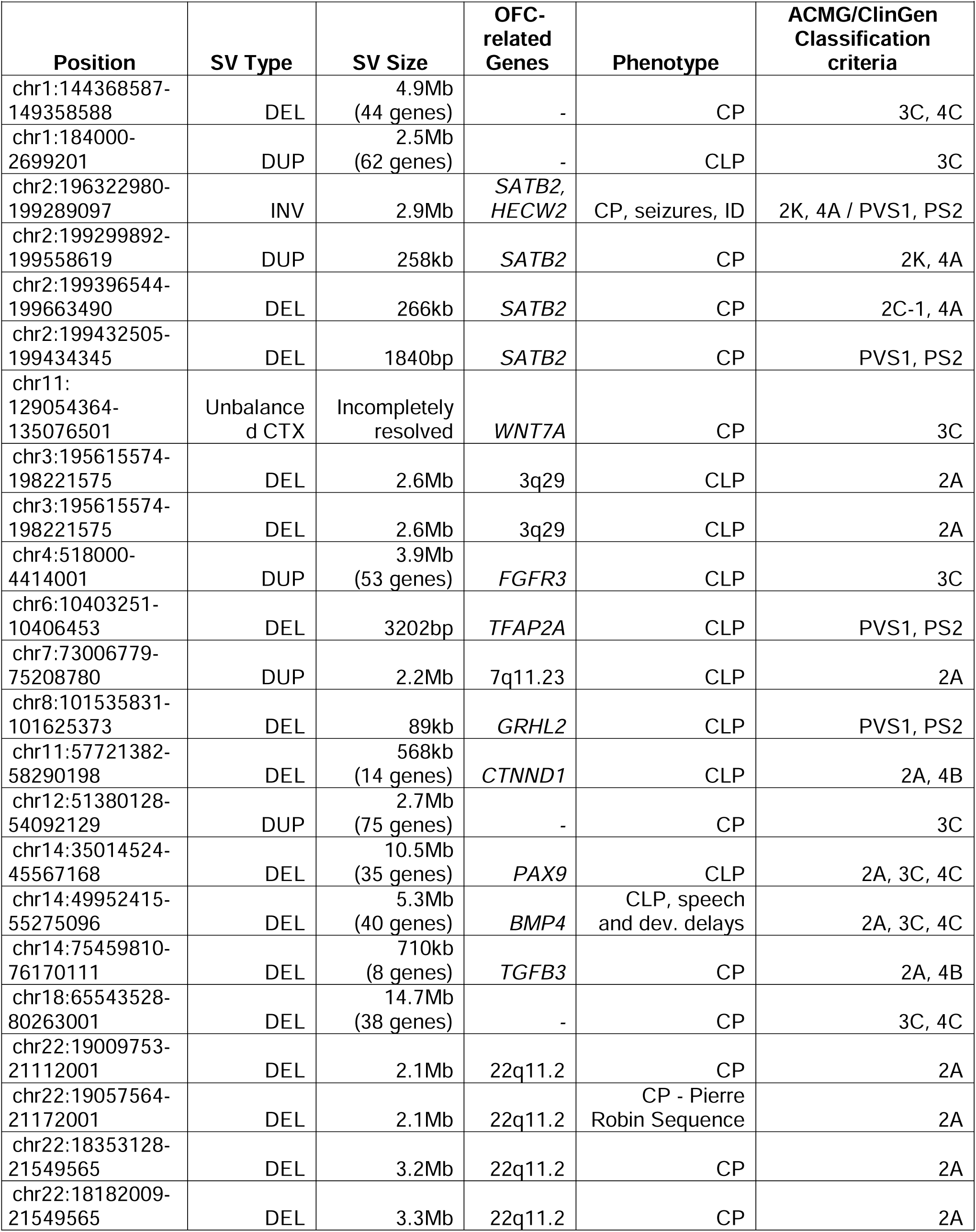
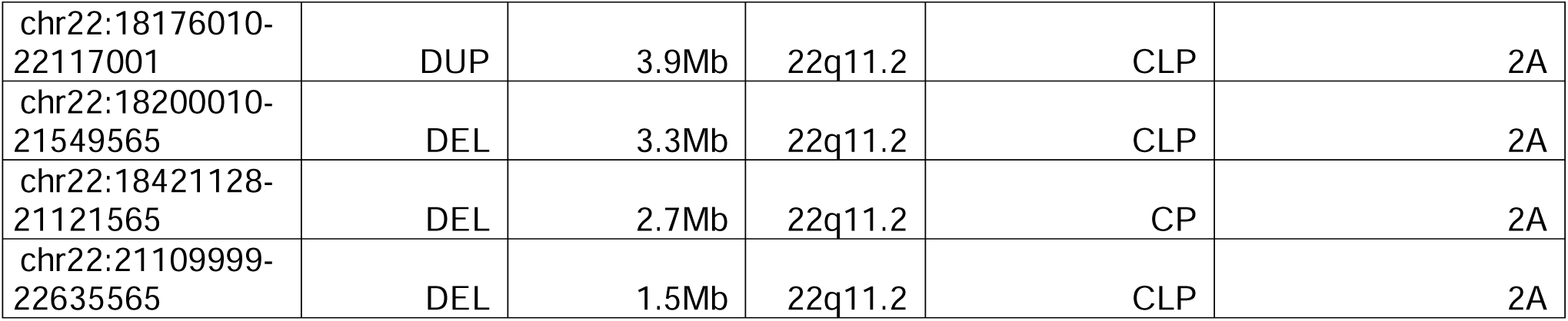
Likely pathogenic dnSVs.

There were 12 SVs exceeding 1Mb, ranging in size from 1.8Mb to 14.7Mb, including 5 duplications, 5 deletions, 1 inversion, and 1 unbalanced translocation (FC 1). Although size alone is not a sufficient determinant for causality, 4 of the 5 deletions result in the loss of more than 35 genes, which is sufficient for calling a variant likely pathogenic (LP) according to the 2015 ACMG-ClinGen recommendations for copy-loss variants^61^. Similarly, 3 of the 5 duplications result in gain of at least 50 genes, meeting the criteria to be deemed LP for copy-gain variants. We note that two of these likely pathogenic variants (14q21.3-q22.3del and 1p36.33dup) overlap genes associated with autosomal dominant OFCs (*BMP4* and *SKI*, *DVL1*, *GNB1*, respectively).

We also consider the 3.0Mb inversion to be pathogenic. It was identified in a proband with CP, seizures, and intellectual disability (ID). Although the recruitment details noted an overdose of anti-epileptic medications as the possible cause, the inversion breakpoints disrupt *SATB2* and *HECW2* (Figure 3). *SATB2*-associated syndrome (MIM: 612313) is a well-established cause of CP, and encompasses other phenotypes such as developmental delays, hypotonia, and seizures^68^. Heterozygous variants in *HECW2* (MIM:617268) cause a syndrome with overlapping clinical features, except for CP^69^. Thus, this inversion offers a complete genetic explanation for the proband’s OFC, seizures, and ID, but the possibility of the environmental exposure being a contributing or modifying factor remains. In total, 9/12 of the large variants can be classified as likely pathogenic, accounting for 0.6% of all sequenced probands.

**Figure 3.**
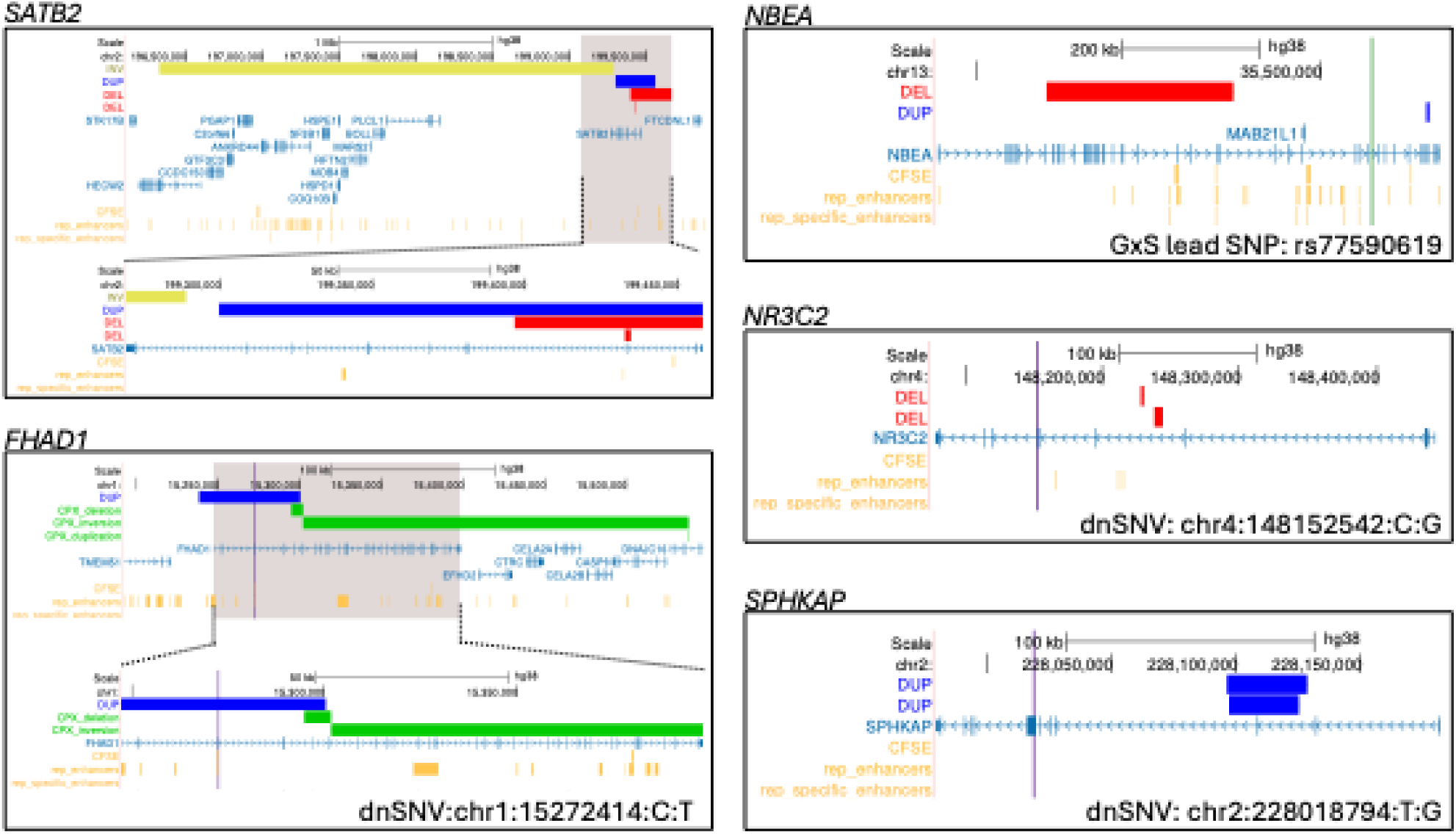
UCSC Genome Browser GRCh38 view of genes implicated by overlapping or recurrent dnSVs, dnSNVs, or common variant signals. (*We can rotate to make this bigger in the paper*). Gray highlights indicate gene boundaries for *SATB2* and *FHAD1*. Variants in red are deletions, blue are duplications, and yellow are inversions. Purple single highlights represent dnSNVs. Green single highlights represent lead SNP from previous gene-by-sex study of orofacial clefts. The enhancer tracks in orange are from the Reproducible Craniofacial ChromHMM Segmentations^50^.

We identified 13 dnSVs that directly overlapped coding regions of 418 OFC-associated genes using a curated set from clinical sequencing panels^70^ (FC 2A). Six of these dnSVs (5 deletions, 1 duplication) disrupt genes associated with autosomal dominant (AD) OFCs and account for 0.4% of cases (Table 1). Among these were 3 additional variants affecting *SATB2* (2 deletions, 1 duplication), bringing the total for *SATB2* to 4 dnSVs (Figure 3), all of which were found in probands with CP. We also identified a 424kb deletion of the last 3 exons of *PDGFC*. Although this gene is less well established in humans (and has not been curated by ClinGen or by NHS Genomic Medicine Service), there are case reports of *de novo* deletions^71^ and missense variants^72^ in *PDGFC* associated with OFCs, and knockout of *Pdgfc* in mice causes a cleft palate^73, 74^.

Given the increased recognition and interest in multiple molecular diagnoses, we also considered whether any individuals in the cohort had multiple SVs that could explain their OFCs, especially among individuals with GDRs, which are incompletely penetrant. Of the 43 individuals with multiple dnSVs (range 2-11 dnSVs per person), 7 had a likely pathogenic SV (GDR or other pathogenic SV) and one or more additional SVs (Supplementary Table 4). Only 1/7 had multiple dnSVs we considered likely pathogenic. This individual had a 2.7Mb deletion of 22q11.2 and 710kb deletion of 14q24.3 that deleted or truncated 8 genes (*BATF*, *ERG28*, *FLVCR2*, *GPATCH2L*, *IFT43*, *JDP2*, *TGFB3*, *TTLL5).* Interestingly, this individual also had two *de novo* missense variants (*HBS1L* and *SFR1*). Although a 22q11.2 deletion alone is sufficient to cause CP^75^, heterozygous variants in *TGFB3* are associated with Loeys-Dietz syndrome 5 (MIM: 615582) and isolated OFCs^76, 77^. Complete loss of *HBS1L* has been reported in a person with multiple anomalies, including submucous CP^78^, and *Hsb1l* knockout mice develop CP^79^. Together, this individual has multiple risk factors for OFC, extending monogenic risk into a heterogeneous and multifactorial etiology. In total, we considered 26 probands to harbor one or more likely pathogenic SVs; therefore, these dnSVs account for 1.67% of the cohort.

### SVs identify new candidate genes

To identify new candidate genes for OFCs, we looked for genes and genetic loci that were 1) impacted more than once by dnSVs (*i.e.,* recurrent SVs) or 2) also harbored a dnSNV identified in prior studies of these cohorts^15, 35^ (Table 2).

**Table 2:** Variants with plausible mechanisms for OFCs.

| Position | SV Type | Length | Evidence for association |
| --- | --- | --- | --- |
| chr2:67887619-67925620 | DEL | 38kb | Deletion of multiple craniofacial enhancers - within range of *MEIS1* |
| chr2:228096618-228128619 | DUP | 32kb | Three de novo events in *SPHKAP* (two dnSVs, dnSNV) |
| chr2:228097618-228125619 | DUP | 28kb | Three de novo events in *SPHKAP* (two dnSVs, dnSNV) |
| chr2:5946332-5947624 | DEL | 1292bp | Deletion of enhancer - within range of *SOX11* |
| chr4:156356665-156780752 | DEL | 424kb | Deletion of last three exons of *PDGFC*; mouse knockout results in CP<sup>73, 74</sup>; case reports of OFC in humans<sup>71</sup> |
| chr4:148237635-148243803 | DEL | 6168bp | Three de novo events in *NR3C2* (two dnSVs, dnSNV) |
| chr4:148228001-148230365 | DEL | 2364bp | Three de novo events in *NR3C2* (two dnSVs, dnSNV) |
| chr6:151641293-154864103 | DEL | 3.2Mb | Large deletion of 10 genes, multiple craniofacial enhancers; potential for regulatory disruption |
| chr6:4255205-4394914 | DEL | 140kb | Deletion of multiple enhancers; near *CDYL*<sup>98</sup> |
| chr7:31027706-31028244 | DEL | 538bp | Deletion within an enhancer; near *MINDY4* (harbors a dnSNV) |
| chr8:140543850-140545050 | DEL | 1200bp | Deletion of exon 14 in *AGO2* |
| chr11:129054364-135076501 | DEL | 6.0Mb | Deletion of 23 genes and multiple craniofacial enhancers |
| chr12:124034831-124049505 | DEL | 14kb | Deletion of multiple craniofacial enhancers; near *NCOR2*<sup>111</sup> |
| chr13:35101313-35372832 | DEL | 271kb | Two dnSV events in *NBEA* |
| chr13:35654081-35657797 | DUP | 3716bp | Two dnSV events in *NBEA* |
| chr16:76777754-76777805 | INS:ME:ALU | 51bp | Insertion into a craniofacial super enhancer |

Excluding large SVs and genomic disordered regions, we identified 8 genes/loci with directly overlapping SVs. SATB2 was overlapped by four dnSVs and each of the following genes was overlapped twice: *FHAD1*, *ZNF722P*, *DAB1*, *PRR20A*, *PTGR1, ZNF483*, *SPHKAP*, and *RPH3A.* Among these genes, only *SATB2*, *FHAD1*, and *SPHKAP* had dnSVs overlapping exons (Figure 3), whereas the other genes are identified as the nearest TSS to the overlapping dnSVs. We also found two non-overlapping intronic deletions in *NR3C2* and two non-overlapping SVs *NBEA*: a deletion of exons 12-38 and a duplication of exons 54 and 55.

Twenty-five dnSVs were annotated to genes in which we also identified dnSNVs in the same cohort (FC 2B, Supplemental Table 2); however, because the majority of the SVs were intergenic (where ‘shared’ genes were found via nearest TSS) or intronic, it is difficult to predict a functional impact. In addition to *SATB2*, *FHAD1*, *SPHKAP*, and *NR3C2,* each of which had a dnSV and one or more protein-altering dnSNVs, there were four genes were implicated by deletions of all or part of their exonic sequence: *GRHL2*, *AGO2*, *NRF1*, and *RANBP2*. We recently described *GRHL2* as associated with OFCs^35^ and we considered this SV pathogenic.

### Regulatory effects of dnSVs

We then considered non-coding dnSVs with potential regulatory roles affecting risk for OFCs. Using a set of 94,534 putative human craniofacial enhancers derived from epigenetic profiling of human embryonic craniofacial tissue during lip and palate development ^50, 80, 81^, we prioritized SVs overlapping any enhancer within 1Mb of an OFC panel gene (FC 3A). In total, there were 14 dnSVs meeting these criteria but as expected with large variants, several of these overlapped both coding and non-coding regions and qualified for multiple classifications. As such, if a variant’s predicted impact better aligned to a higher priority category (*e.g*., overlap with coding regions of OFC panel genes), we took that interpretation. After this filtering, there were 2 variants remaining.

One dnSV was a 222bp duplication in a craniofacial enhancer 400kb 5’ to *IRF6,* which has a well-established association with both syndromic^82^ and non-syndromic forms^83–86^ of OFCs. The duplication is within the *IRF6* topologically associated domain (TAD) in embryonic stem cells (ESCs)^51^ but not cranial neural crest cells (CNCCs)^49, 50^, though HiC data^87^ show *IRF6* is within range (Supplemental Figure 3). Taken altogether, these data suggest potential cell-specific interactions or effects for this enhancer. Also in this region was a 1,689bp deletion of exon 9 of neighboring gene *TRAF3IP3* (Supplemental Figure 4). While this deletion does not specifically overlap an enhancer (and is not included in our listed counts), it does overlap a locus previously identified by GWAS^88^. In that study, the *TRAF3IP3* peak (lead SNP rs2235370-C-A) was found to be independent of a second peak mapped to *IRF6*. The mechanism by which *TRAF3IP3* or this locus mediates OFC risk is unclear, but our data further supports this *TRAF3IP3* locus as being relevant.

To identify novel candidate regions, we looked for dnSVs overlapping craniofacial enhancers near genes with protein-altering dnSNVs. There were two dnSVs found within 1Mb of at least one gene with a dnSNV^15, 35^ (FC 3B). These included a 538bp deletion ∼130kb downstream of *MINDY4* and an 87bp deletion ∼150kb upstream of *CTNNA1*. Neither gene has been associated with OFCs to date; however, *CTNNA1* is a member of the cadherin family, some of which have been implicated in OFCs, including *CTNND1*^89^ and *CTNNA2*^13, 90, 91^. It is also ubiquitously expressed in developing craniofacial tissue^92^, making *CTNNA1* another potential candidate for OFCs.

We also considered the remaining variants that overlapped enhancers (FC 3C) on an individual basis, finding three more variants of interest. The first is a 1,292bp deletion in 2p25.2 that overlaps a highly conserved enhancer region. Based on Hi-C data, this enhancer seems to interact with *SOX11* (Figure 4), which is associated with Coffin Siris syndrome 9 (MIM:615866) which can include OFCs^93, 94^. Similarly, there is a 38kb deletion in 2p14 that overlaps an enhancer which appears to interact with *MEIS1* (Supplemental Figure 5). *MEIS1* has been less implicated in craniofacial biology than its family member *MEIS2*^95, 96^, but it has been shown to play a role in craniofacial morphogenesis in zebrafish^97^. Lastly was a 139kb deletion that overlapped multiple enhancers in 6p25.1 (Supplemental Figure 6). A plausible target of these enhancers is *CDYL,* which has been reported as significantly differentially methylated in discordant monozygotic twins with Van der Woude syndrome^98^ (MIM: 119300).

**Figure 4:**
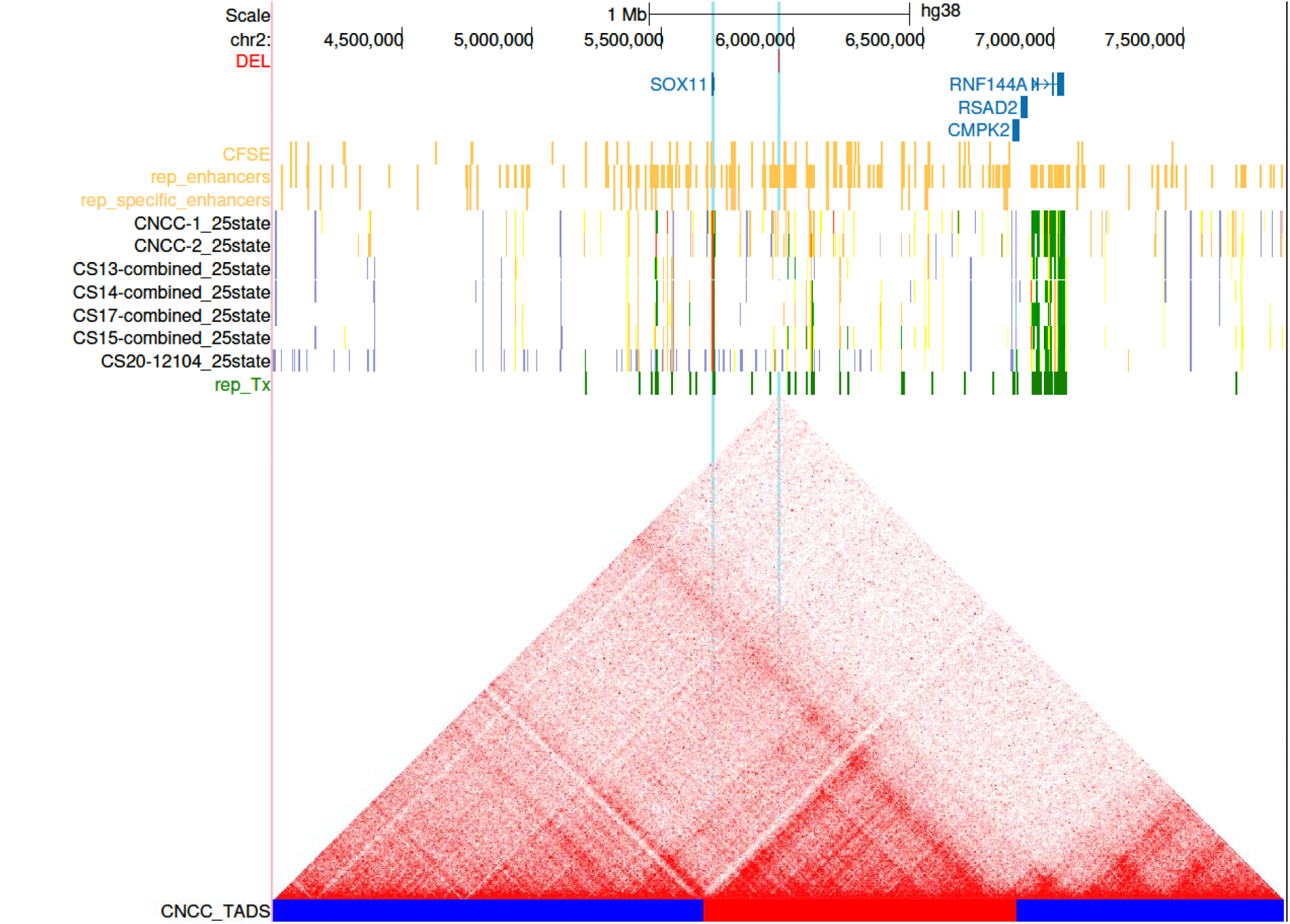
UCSC Genome Browser GRCh38 view of a dnSV deletion of an enhancer near *SOX11*. The identified deletion and *SOX11* are highlighted in light blue. Visible tracks include strong enhancers shown in orange^50^, the 25-state ChromHMM segmentations, and the combined craniofacial Hi-C data^87^. Blue and red bars below Hi-C matrix indicate distinct TADs.

To further investigate the 16 genes identified from our analyses of multiple/overlapping dnSVs, dnSNVs, or regulatory effects, we examined expression patterns from publicly available snRNA-seq data in human embryonic craniofacial tissue. We found that these genes were co-expressed most strongly in ectodermal cell types and in mesenchymal cell types corresponding to the palate (Figure 5A). The cell subtypes expressing these candidate genes also have marker genes that are enriched for dnSNVs from prior analyses; this convergence of evidence supports that, in aggregate, some of these SVs may contribute to OFC risk. Individually, the candidate genes showed highly variable expression patterns, with some (*e.g., FHAD1*, *DDX27*, *SPHKAP*) showing expression restricted to a subset of cells and cell types and others (*e.g., NBEA*) showing moderate expression throughout a range of cell types (Figure 5B). Collectively, these data provide evidence that many of genes nominated by analysis of dnSVs are expressed in the right time and place to be relevant to pathogenesis of OFCs. The absence of a unifying expression pattern is consistent with the heterogeneity of OFC risk genes and the numerous biological processes required for proper craniofacial development.

**Figure 5.**
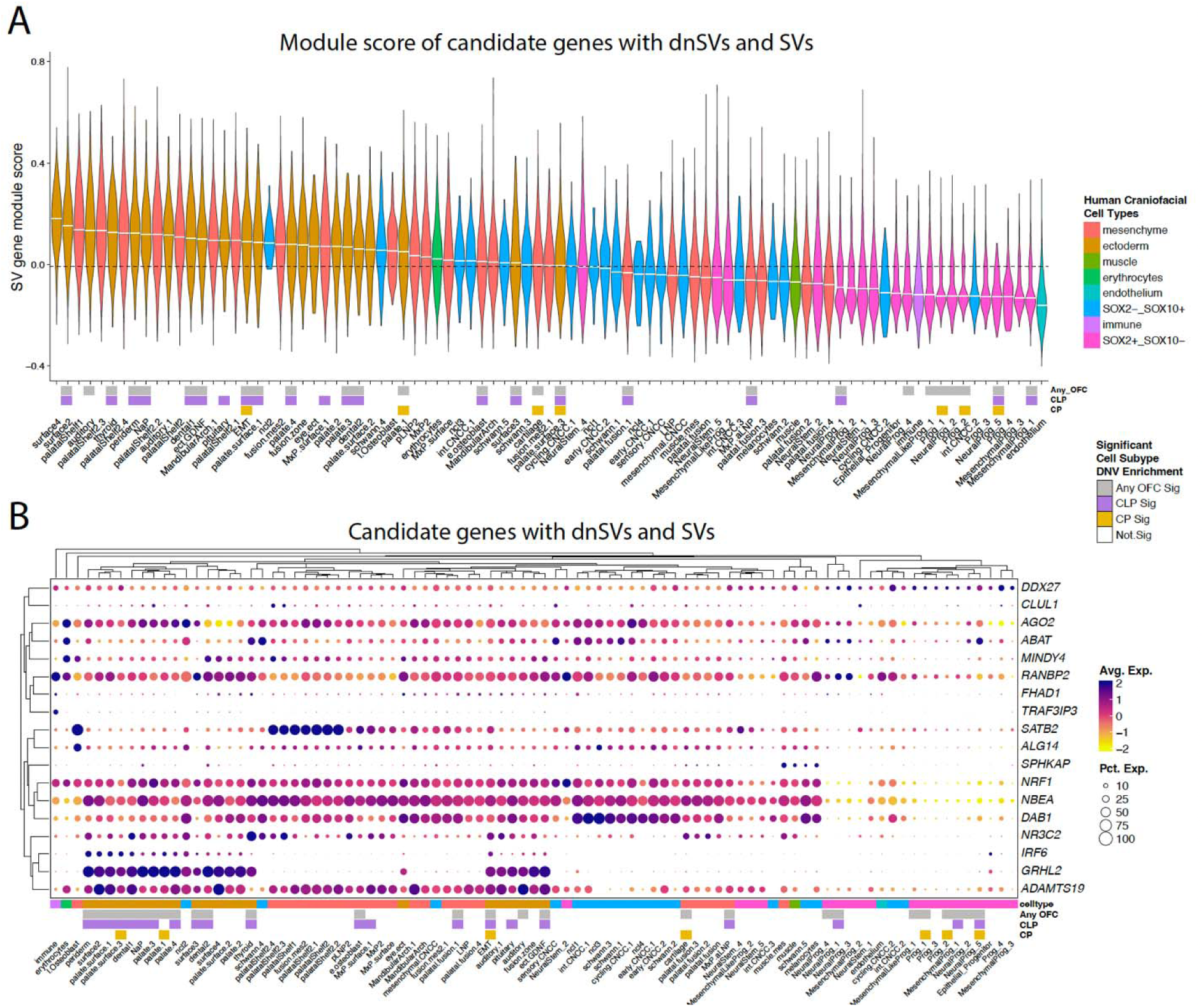
Expression patterns of candidate genes with dnSVs and SVs in human embryonic facial tissue. (A) Violin plot for all cells of a given subtype from Khouri-Farah *et al.* 2026^1^ based on a module score calculated from candidate genes with dnSVs and SVs in human embryonic facial tissue. (B) Clustered dot plot of average and percent expression for candidate genes with dnSVs and SVs in human embryonic facial tissue across human cell subtypes from Khouri-Farah *et al.* 2026^1^ for candidate genes with dnSVs and SVs. In both panels, significant enrichments across cell subtypes from Khouri-Farah *et al.* 2026^1^ of protein damaging de novo variation in orofacial cleft trios from the Gabriella Miller Kids First program^2,3^ are shown below, colored by OFC subtype. Note that *IRF6* and *SATB2* added to the 16 candidate genes as reference points as established OFC genes.

## Discussion

Here we describe 375 dnSVs in 307 probands from a cohort of 1,549 trios with OFCs. We found that across subtypes, probands with CL had the highest rate of structural variation, followed by CLP, then CP. This was somewhat unexpected, as previous work has found no difference in copy number variation burden between OFC subtypes^30^, and the diagnostic yield has been found to be low in a previous subset of CL (2.8%)^48^. Interestingly, we found the rate of duplications per genome was higher in CL and CLP than in CP, and that duplications in CL had trend toward higher pTriplo scores. This suggests a hypothesis of higher sensitivity to increased gene dosage in CL, which will need to be tested in larger cohorts.

In total, we estimate that dnSVs classified as likely pathogenic account for a diagnostic yield of 1.67% in this cohort. Pathogenic SVs accounted for 2.8% of CP probands and 1.4% of CLP probands but none of the CL probands had a pathogenic SV. An additional 0.96% carry dnSVs that are plausibly relevant to OFC risk but lack sufficient evidence for classification as causal; these represent 1.8% of CL, 0.5% of CLP, and 0.6% of CP probands. Overall, these results are consistent a low diagnostic yield in CL and support the hypothesis that the genetic architecture for CL is different from CLP and CP. A caveat to these findings, however, is an important note that there may under-enrichment for dnSVs that are easily identified with prescreening methods such as microarray. Individuals with known causal SVs may not be ascertained by our study, and thus not represented by this cohort.

Only 1 of the 26 people harboring pathogenic variants was reported with additional phenotypes at the time of recruitment. It remains challenging to determine if: 1) dnSVs are contributing more to isolated OFC etiology than previously considered due to current testing protocols^99^ (*i.e.,* isolated OFCs do not always undergo genetic testing), 2) proband ascertainment during infancy leads to under-reporting of phenotypes that were not apparent at the time of enrollment, or 3) these findings are representative of rare but benign or mild variation that is not modulating OFC risk. While the true explanation is likely a combination of these factors, our data provide evidence that dnSVs are part of the genetic architecture for isolated OFCs. Further, we speculate that these results may even represent a lower contribution by SVs to OFC risk than what the true incidence might be. For example, individuals harboring SVs who have undergone genetic testing such as chromosomal microarray analysis would have been provided positive test results that can decrease the likelihood of enrollment in a research cohort. Thus, the ascertainment of our probands is likely also affecting our overall results.

We also used these dnSVs to identify genes of interest without previously known association with OFCs. Using a combination of dnSVs, dnSNVs, and regulatory information, we suggest *CTNNA1*, *FHAD1*, *NBEA*, *NR3C2*, and *SPHKAP* as the primary genes of interest from this study. There are two reports of cleft palate in individuals with hyperaldosteronism due to exon deletions in *NR3C2*^100, 101^, which encodes the mineralocorticoid receptor. We observed three total *de novo* events in this gene, but both SVs were intronic deletions and the effect on *NR3C2* splicing or expression is unknown. We found an exon 3 duplication in two individuals in *SPHKAP,* which encodes an A-kinase-anchoring protein that plays a role in cell proliferation and apoptosis^102^ and is enriched in brain and heart tissue. We found this gene to be expressed in craniofacial muscle, which may be consistent with its biological function in the heart, but a mechanism for involvement in OFCs is not apparent. The variants in *FHAD1* clearly impacted its exonic sequence but this gene showed relatively low and non-specific expression in craniofacial tissues. All of the variants, including a *de novo* missense variant, also overlapped putative craniofacial enhancers, so we cannot exclude that these variants influence OFC risk through regulation of a nearby gene. Finally, the two SVs in *NBEA* provide additional evidence for this gene be relevant to OFCs. Previous work has found common SNVs in an intron of *NBEA* associated with increased CL/P risk in a sex-specific manner in a genome-wide gene by sex interaction study^103^ and homozygous knockout of *Nbea* in mouse results in cleft palate^104^.

While more challenging to interpret, we also found evidence of dnSVs with potential regulatory effects. Enhancer disruption has been reported as causal for other conditions, including Pierre Robin Sequence^105, 106^, congenital heart defects^107^, and triphalangeal thumb-polysyndactyly syndrome (MIM:190605)^108^. We found both duplications and deletions of craniofacial enhancers which we speculate could affect gene dosage. Although the mechanism by which these loci may confer risk for OFC is unknown, this study provides evidence for their potential involvement. As more is discovered about gene regulation and dysregulation in structural birth defects, interpretation of these kinds of variants can be improved. Alternatively, these variants could be tested further in model systems to better predict their effects.

In the context of prior work on SVs in OFCs, we did not find any dnSVs in the genes or loci previously reported (*e.g., COBLL1*, 7p14)^27, 30, 33, 34^. Although there is a partial sample overlap between this study and past cohorts, this lack of replication not unexpected. Both our and others’ cohorts have relatively small sample sizes, and our study is focused on dnSVs, which occur at a comparatively lower abundance than inherited SVs. Technology is also likely to play an important role as previous work primarily used SNP array data or arrayCGH and we used short-read WGS. Although this method expands SV detection to across the genome, short read WGS cannot confidently call SVs in repetitive regions, resulting in fewer called SVs than might be found with long-read data^109, 110^. Thus, incorporation of long-read sequencing technologies for future studies would be beneficial in resolving SVs in OFCs.

In summary, our study provides a comprehensive overview of dnSVs in people with OFCs and offers evidence that they are a rare but important contributor to its genetic architecture. This study also demonstrates how integration of genetic variants of multiple types and disease-relevant epigenetic and expression data can be used to identify new candidate genes through the interpretation of coding and noncoding variant effects. Although this study offers more hypotheses than confirmed causal relationships, our specific findings and catalogue of dnSVs in these cohorts may be used to foster gene discovery in rare disease and to empower studies on diverse topics from genetics of OFCs and other birth defects to structural variation and its impact on gene regulation.

## Data Availability

All original sequence data analyzed is available from dbGaP and/or the Kids First data portal. All structural variants analyzed in this manuscript are contained in the supplemental data.

## Acknowledgements

We thank the participants and collaborating clinical and recruitment staff for their efforts that made this study possible. Participant recruitment, assembly of phenotypic information, and data analysis were supported by National Institutes of Health (NIH) grants: F31-DE032588 (KR), R35-GM119465 (JC), R01-DE028945 (JC), R01-DE028342 (EJL), R01-DE027983 (EJL), R01/R37-DE008559 (JCM), R01-DE016148 (MLM, SMW); R01-DE032319 (MLM, EJL, SMW, JRS), R01-DE031261 (HB), R01DE011931 (JH). We thank the California Department of Public Health, Maternal Child and Adolescent Division, for providing data for these analyses. This work was supported by the Centers for Disease Control and Prevention, Centers of Excellence No. U01-DD001033.

Sequencing services for CPSeq (X01-HG010835) were provided by the Center for Inherited Disease Research (CIDR). CIDR is fully funded through a federal contract from the NIH to The Johns Hopkins University, contract number HHSN268201700006I. Sequencing services for the Gabriella Miller Kids First cohorts were supported by the NIH: X01-HG010835, X01-HL0132363, X01-HD100701, X01-DE030062, X01-DE032472, X01-HL132363, X01-HL136465.

## Data availability

Sequence and phenotype data is available from the Database of Genotypes and Phenotypes (dbGaP, ncbi.nlm.nih.gov) under study accession #s: phs002220.v1.p1 (CPSeq), phs001168.v2.p2 (GMKF US, European), phs001420.v1.p1(GMKF Colombian), phs000094.v1.p1 (GMKF Taiwanese), phs002595.v1.p1 (GMKF Philippines)

