## Supplemental Data for "Characterization of *de novo* structural variants in trios with orofacial clefts"

**Supplementary Figures and Tables**

| **Supplementary Table 1: Description of OFC probands (N, % of cohort)** | | | | |
| --- | --- | --- | --- | --- |
|  | CL | CLP | CP | Total |
| Male | 124 (8.0%) | 596 (38.5%) | 189 (12.2%) | 909 (58.6%) |
| Female | 91 (5.9%) | 288 (18.6%) | 261 (16.8%) | 640 (41.3%) |
| Syndromic | 3 (0.2%) | 19 (1.2%) | 32 (2.1%) | 54 (3.5%) |
| Nonsyndromic | 86 (5.6%) | 480 (31.0%) | 413 (26.7%) | 979 (63.2%) |
| Status unknown | 126 (8.1%) | 385 (24.9%) | 5 (0.3%) | 516 (33.3%) |
| Total (%) | 215 (13.9%) | 884 (57.1%) | 450 (29.1%) | 1549 (100%) |

**Supplementary Table 2. All dnSNVs (xlsx)**

**Supplementary Table 3. All dnSVs (xlsx)**

| **Supplementary Table 4. Individuals with multiple dnSVs** | | | | | | | | | | | |
| --- | --- | --- | --- | --- | --- | --- | --- | --- | --- | --- | --- |
| Individual | Total SV count | Counts | | | | | | | | | |
|  |  | DEL | INS:ME:  LINE1 | DUP | INS:ME:  ALU | INS:ME:  SVA | INS | INV | CPX | CTX | GDR |
| PA1828 | 11 | 0 | 0 | 11 | 0 | 0 | 0 | 0 | 0 | 0 | 0 |
| GMKFPHIndiv0154 | 5 | 0 | 0 | 5 | 0 | 0 | 0 | 0 | 0 | 0 | 0 |
| GMKFPHIndiv0892 | 5 | 0 | 0 | 5 | 0 | 0 | 0 | 0 | 0 | 0 | 0 |
| IA3040 | 4 | 0 | 0 | 4 | 0 | 0 | 0 | 0 | 0 | 0 | 0 |
| GMKFPHIndiv0001 | 3 | 1 | 0 | 2 | 0 | 0 | 0 | 0 | 0 | 0 | 0 |
| GMKFPHIndiv0493 | 3 | 0 | 0 | 3 | 0 | 0 | 0 | 0 | 0 | 0 | 0 |
| GMKFPHIndiv0709 | 3 | 0 | 0 | 3 | 0 | 0 | 0 | 0 | 0 | 0 | 0 |
| GMKFPHIndiv0937 | 3 | 0 | 0 | 2 | 1 | 0 | 0 | 0 | 0 | 0 | 0 |
| 202030530 | 3 | 1 | 0 | 0 | 1 | 0 | 0 | 0 | 0 | 0 | 1 |
| 202030205 | 3 | 0 | 0 | 3 | 0 | 0 | 0 | 0 | 0 | 0 | 0 |
| GMKFC33630 | 3 | 3 | 0 | 0 | 0 | 0 | 0 | 0 | 0 | 0 | 0 |
| IA2671 | 3 | 0 | 0 | 3 | 0 | 0 | 0 | 0 | 0 | 0 | 0 |
| GMKFPHIndiv0667 | 2 | 1 | 0 | 0 | 0 | 0 | 1 | 0 | 0 | 0 | 0 |
| GMKFPHIndiv0682 | 2 | 0 | 0 | 0 | 2 | 0 | 0 | 0 | 0 | 0 | 0 |
| GMKFPHIndiv1086 | 2 | 2 | 0 | 0 | 0 | 0 | 0 | 0 | 0 | 0 | 0 |
| 201900125 | 2 | 0 | 0 | 2 | 0 | 0 | 0 | 0 | 0 | 0 | 0 |
| 201904436 | 2 | 1 | 0 | 0 | 0 | 0 | 0 | 0 | 0 | 0 | 1 |
| 202030542 | 2 | 2 | 0 | 0 | 0 | 0 | 0 | 0 | 0 | 0 | 0 |
| 202030031 | 2 | 1 | 0 | 1 | 0 | 0 | 0 | 0 | 0 | 0 | 0 |
| 202030262 | 2 | 2 | 0 | 0 | 0 | 0 | 0 | 0 | 0 | 0 | 0 |
| 202030106 | 2 | 1 | 0 | 0 | 0 | 0 | 0 | 1 | 0 | 0 | 0 |
| 202030052 | 2 | 0 | 0 | 1 | 0 | 0 | 1 | 0 | 0 | 0 | 0 |
| 202030940 | 2 | 1 | 0 | 0 | 1 | 0 | 0 | 0 | 0 | 0 | 0 |
| GMKFC15945 | 2 | 1 | 0 | 1 | 0 | 0 | 0 | 0 | 0 | 0 | 0 |
| IA1144 | 2 | 1 | 0 | 0 | 1 | 0 | 0 | 0 | 0 | 0 | 0 |
| IA2854 | 2 | 2 | 0 | 0 | 0 | 0 | 0 | 0 | 0 | 0 | 0 |
| KF19234_01 | 2 | 1 | 0 | 0 | 1 | 0 | 0 | 0 | 0 | 0 | 0 |
| MD0208 | 2 | 0 | 0 | 1 | 0 | 0 | 0 | 0 | 0 | 0 | 1 |
| MD0214 | 2 | 0 | 0 | 2 | 0 | 0 | 0 | 0 | 0 | 0 | 0 |
| MD0301 | 2 | 0 | 0 | 2 | 0 | 0 | 0 | 0 | 0 | 0 | 0 |
| MD0310 | 2 | 0 | 0 | 2 | 0 | 0 | 0 | 0 | 0 | 0 | 0 |
| 201904633 | 2 | 2 | 0 | 0 | 0 | 0 | 0 | 0 | 0 | 0 | 0 |
| PA1645 | 2 | 0 | 0 | 2 | 0 | 0 | 0 | 0 | 0 | 0 | 0 |
| PA2065 | 2 | 0 | 0 | 2 | 0 | 0 | 0 | 0 | 0 | 0 | 0 |
| PA2212 | 2 | 0 | 0 | 1 | 0 | 0 | 0 | 0 | 1 | 0 | 0 |
| PA2308 | 2 | 0 | 0 | 1 | 1 | 0 | 0 | 0 | 0 | 0 | 0 |
| GMKFPHIndiv0100 | 2 | 0 | 0 | 1 | 0 | 0 | 0 | 0 | 1 | 0 | 0 |
| GMKFC33569 | 2 | 2 | 0 | 0 | 0 | 0 | 0 | 0 | 0 | 0 | 0 |
| GMKFPHIndiv1087 | 2 | 0 | 0 | 1 | 0 | 0 | 0 | 0 | 0 | 0 | 1 |
| PA1765 | 2 | 0 | 0 | 1 | 0 | 0 | 0 | 0 | 0 | 0 | 1 |
| 202030241 | 2 | 0 | 0 | 2 | 0 | 0 | 0 | 0 | 0 | 0 | 0 |
| 202031141 | 2 | 1 | 0 | 1 | 0 | 0 | 0 | 0 | 0 | 0 | 0 |
| IA3025 | 2 | 0 | 0 | 0 | 0 | 0 | 0 | 0 | 2 | 0 | 0 |
| Has dnSV/GDR variant considered pathogenic  Has GDR variant with incomplete penetrance – unknown effect in this proband  Number of variants per category | | | | | | | | | | | |

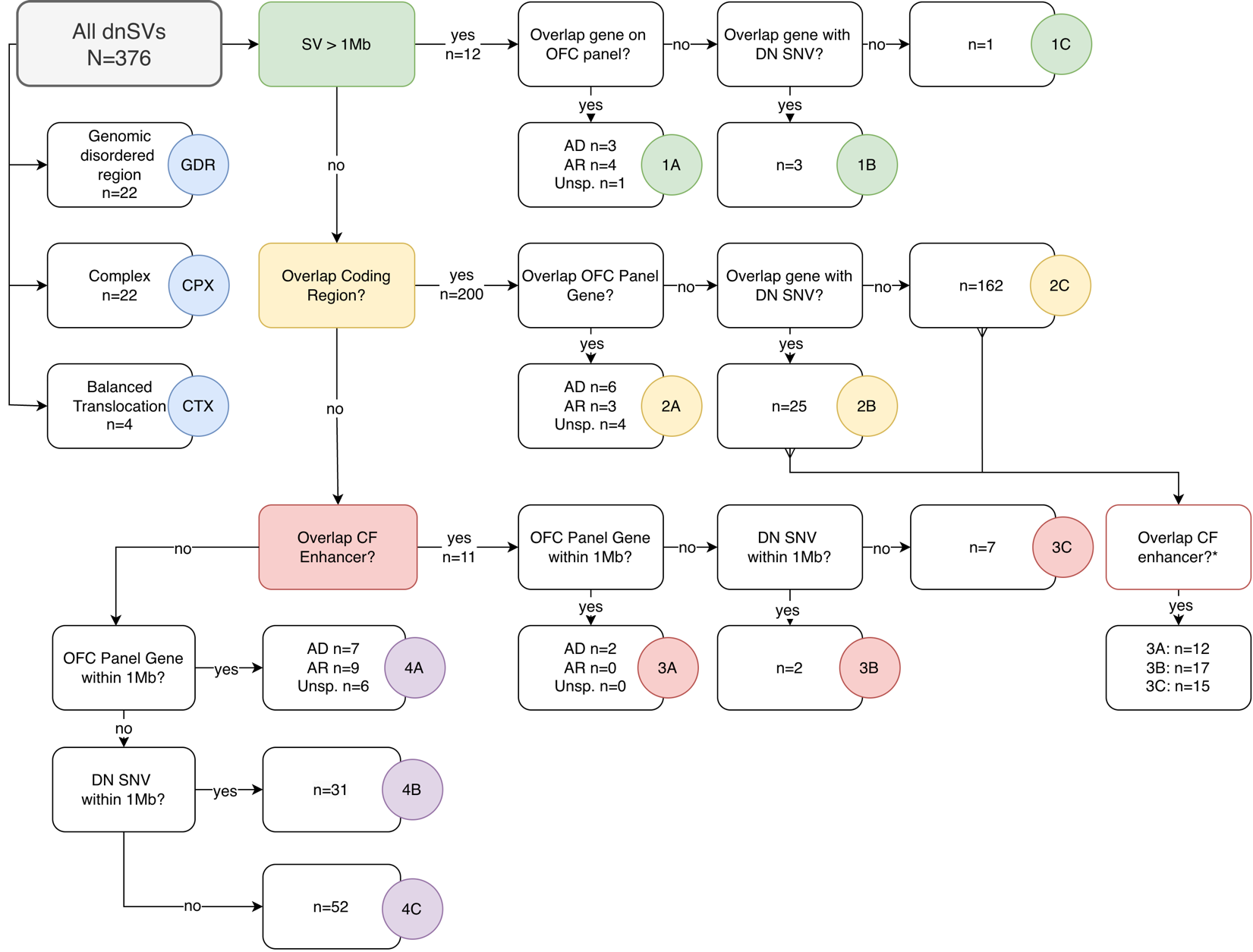

**Supplemental Figure 1. Flowchart for categorizing and interpreting dnSVs**

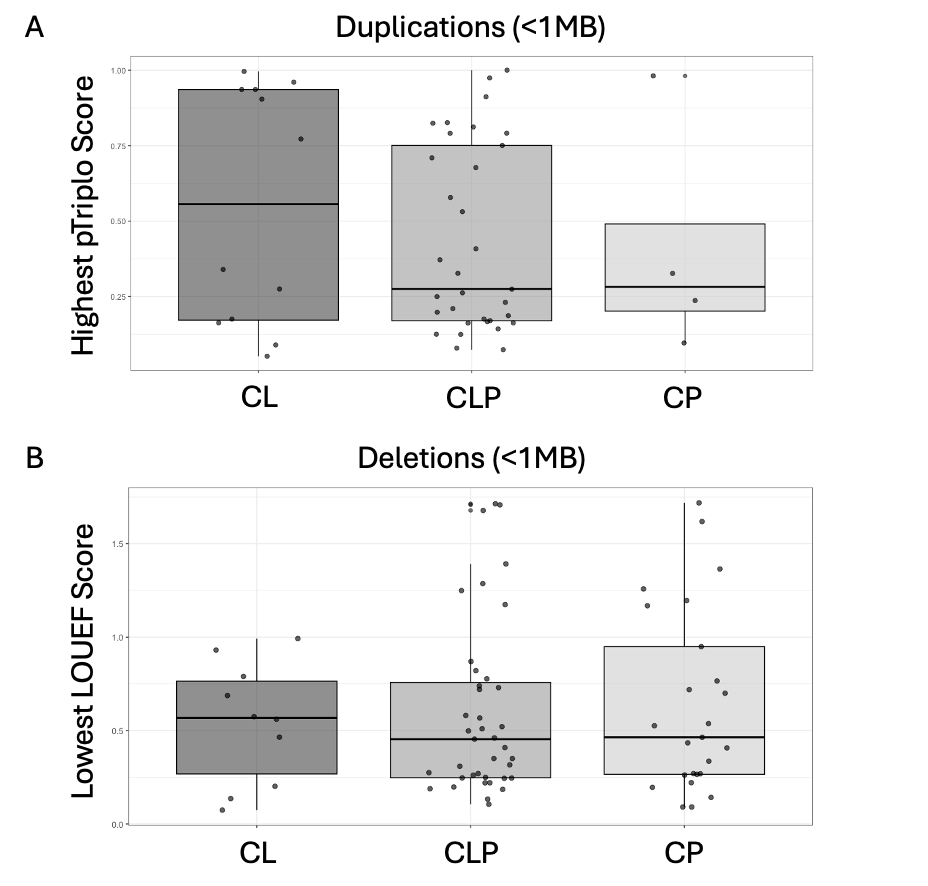

**Supplemental Figure 2: Box plots showing dosage sensitivity scores** for the A) highest pTriplo score of all genes within a duplication or B) lowest LOEUF score of all genes within a deletion.

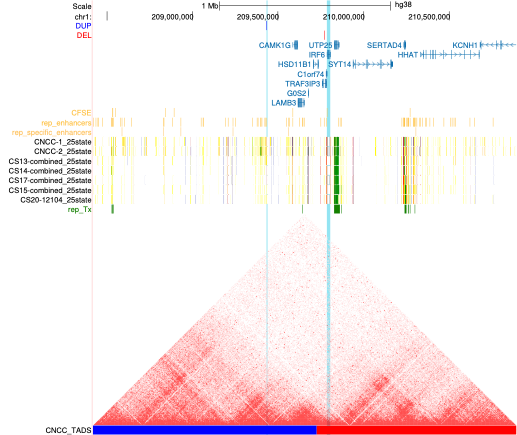

**Supplemental Figure 3: UCSC Genome Browser GRCh38 view of IRF6 and the surrounding region.** The identified duplication and *IRF6* are highlighted in light blue. Visible tracks include strong enhancers shown in orange^47^, the 25-state ChromHMM segmentations, and the combined craniofacial Hi-C data^84^.

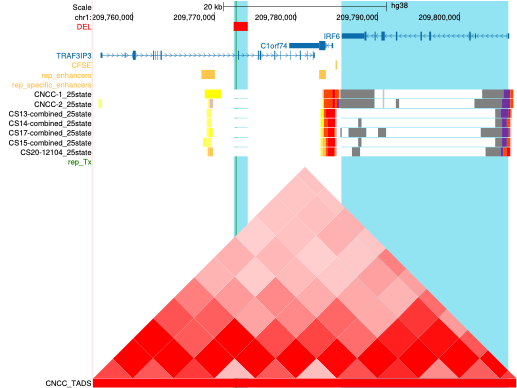

**Supplemental Figure 4: UCSC Genome Browser GRCh38 view of IRF6 and nearby region.** The blue highlighted regions are *IRF6* and the dnSV deletion, and the green highlight represents a previously identified GWAS signal (rsID). Visible tracks include strong enhancers shown in orange^47^, the 25-state ChromHMM segmentations, and the combined craniofacial Hi-C data^84^.

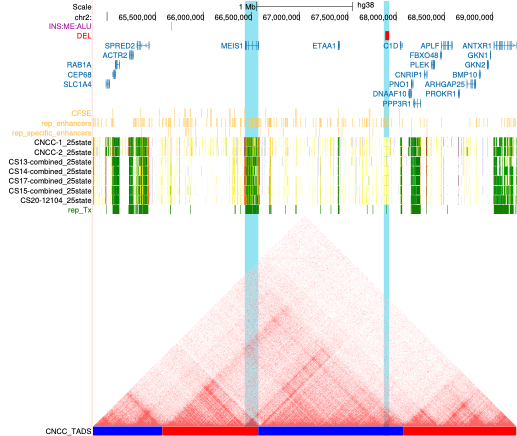

**Supplemental Figure 5: UCSC Genome Browser GRCh38 view of *MEIS1* and the surrounding region.** Blue highlighted regions are *MEIS1* and deletion of an enhancer within range. Visible tracks include strong enhancers shown in orange^47^, the 25-state ChromHMM segmentations, and the combined craniofacial Hi-C data^84^.

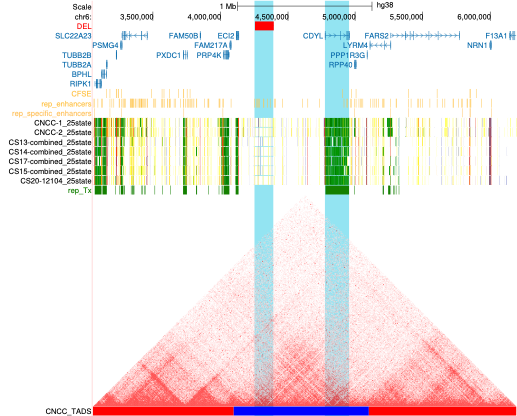

**Supplemental Figure 6: UCSC Genome Browser GRCh38 view of a dnSV deletion of several enhancers and the surrounding genes.** Blue highlighted regions are *CDYL* and deletion of multiple enhancers within range. Visible tracks include strong enhancers shown in orange^47^, the 25-state ChromHMM segmentations, and the combined craniofacial Hi-C data^84^.

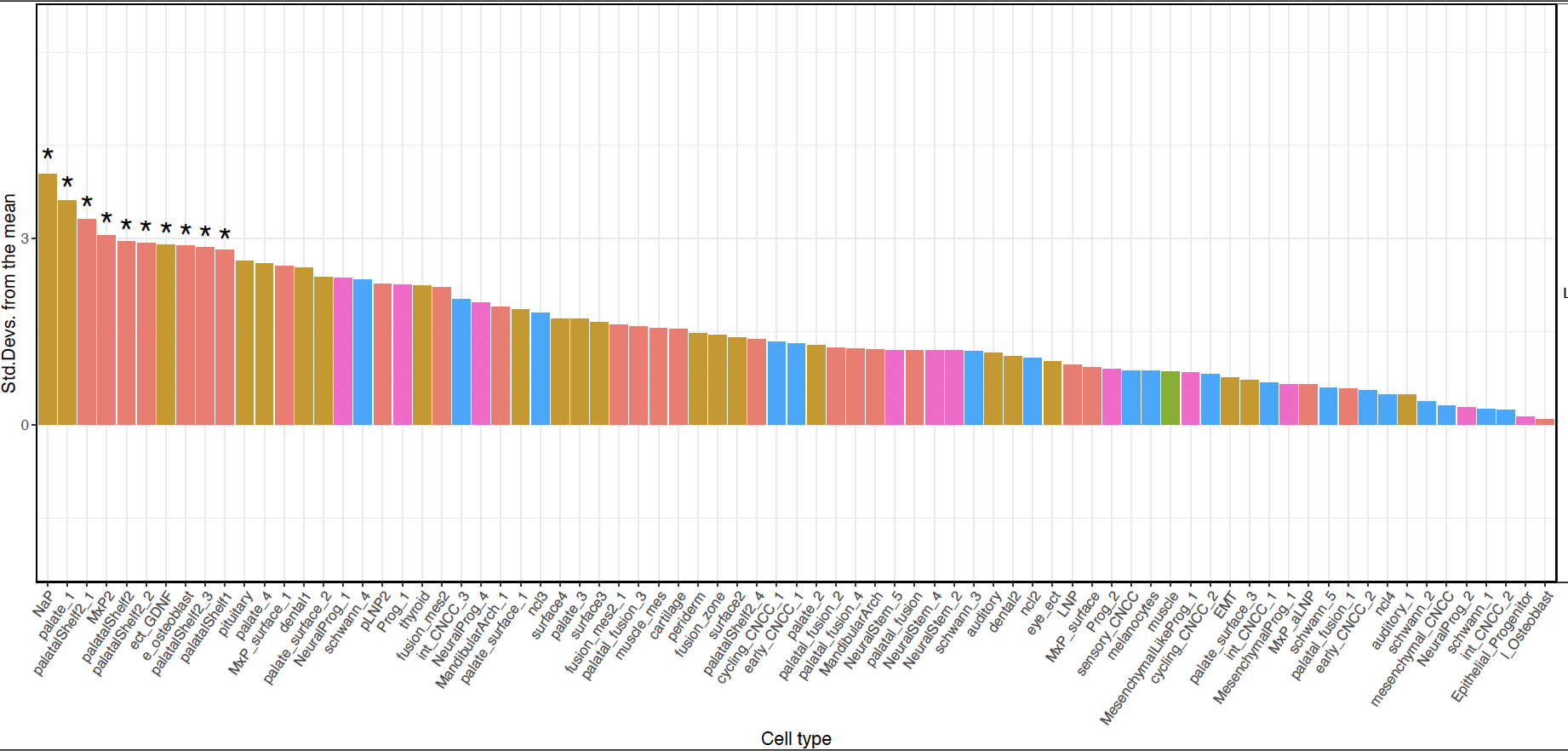

**Supplemental Figure 7.** Bar plot of standard deviations from the mean of bootstrapping tests performed by EWCE^4^ method for all genes with protein-altering dnSNVs across human craniofacial cell subtypes. Asterisks indicate significant subtype enrichments (Benjamini-Hochberg corrected p-value < 0.05).
